# Fisher information matrix computation for joint longitudinal and survival models to support clinical study design and covariate effect assessment

**DOI:** 10.64898/2026.05.28.26354340

**Authors:** Lucie Fayette, Karl Brendel, France Mentré

**Author notes:** **Correspondence:** Lucie Fayette.

## Abstract

Joint modelling of longitudinal data using non-linear mixed effects models and time-to-event outcomes provides a suitable framework to account for informative censoring when estimating biomarker dynamics and quantifying event risk using covariates and longitudinal trajectories. Their usefulness in clinical research depends on data collection design, particularly to precisely estimate the association (link) parameter between longitudinal and survival processes. However, optimal design strategies have so far been addressed separately for longitudinal and survival endpoints and remain unexplored for joint models. We propose two Fisher Information Matrix (FIM) computation methods for joint models, relying on Monte-Carlo integration over observations combined with either Markov Chains Monte-Carlo or Adaptive Gaussian Quadrature to integrate random effects. Their accuracy is assessed against clinical trial simulations in an oncological example based on the HORIZON III study with a tumour-growth–survival model including discrete and continuous covariates. We apply these methods to quantify the impact of follow-up duration, sampling richness, sample size, and covariate distribution on parameter uncertainty and test power.

In our example, longitudinal-parameter uncertainty is barely affected by follow-up duration or sampling richness, whereas survival-parameter uncertainty decreases substantially from 1-year to 2-year follow-up. The number of subjects needed (NSN) to achieve <15% uncertainty on the link parameter is comparable for a 2-year rich design and a 3-year sparse design. Optimal covariate distributions are stable across designs and systematically improve test power, outperforming longer and richer but non-optimised designs.

These FIM-based methods accurately predict uncertainty and test powers, enabling design evaluation and NSN computation for joint-model-based clinical studies.

## 1 Introduction

Joint modelling of longitudinal data and time-to-event outcomes has gained increasing attention in medical research, as it enables the simultaneous analysis of biomarkers measured repeatedly over time and an event-time outcome such as death. The objective of such analyses is often to identify biomarkers with strong prognostic value for the event time [1]. In joint models, the longitudinal biomarker trajectory is typically described using a mixed-effects model, while the time-to-event process is modelled using a survival model [1]. These models provide a suitable framework to estimate biomarker evolution without bias from informative censoring, assess event risk using both covariates and longitudinal information, and investigate the relationship linking the longitudinal and survival processes [2]. Recent applications include the investigation of the association between tumour size and survival after treatment initiation in oncology [3, 4, 5], between a daily Sequential Organ Failure Assessment score measurements in intensive care units (ICU) and competing risks of ICU death and discharge [6], and between viral load and death in patients with SARS-CoV-2 [7]. In pharmacology and pharmacometrics, biomarker trajectory is captured through non-linear mixed effects models (NLMEM) [8].

While joint models are increasingly used, their practical application in clinical studies depends on the design of the data collection scheme. Especially, the number of subjects, the frequency and timing of measurements, the distribution of covariates across individuals, may influence the amount of information available to estimate both biomarker dynamics and their association with the event. For drug development, regulatory agencies [9] recommend both clinical trial simulation (CTS) and optimal design strategies. While CTS remains the gold standard for final design validation, it becomes prohibitive when exploring large design spaces, optimising covariate distributions, or evaluating multiple trade-offs between follow-up duration, sampling richness, and sample size. FIM-based methods uniquely allow such systematic and efficient exploration. Optimal design theory relies on the Fisher Information Matrix (FIM) [10], which inverse, according to the inequality of Rao-Cramer, is a lower bound for the variance of any unbiased estimator. Therefore, expected standard errors (SE) resulting from a design can be derived.

In the NLMEM framework, the FIM has no closed-form solution and requires the computation of two intractable integrals, respectively with respect to the random effects and to the observations. For continuous response with Gaussian error, first-order linearization of the NLMEM yields a Gaussian approximation with an analytical FIM [11, 12], which has been shown to be appropriate for design evaluation and optimisation [13], and is implemented in various software tools [14].

Design strategies for survival outcomes have also been explored in settings where the FIM has a closed form [15] and have been shown to be efficient to improve parameter estimations and design more ethical and timely studies. It was also shown that accounting for covariates can significantly change the optimal design in terms of sample size and number of measurements for each subject, and that misspecification of either covariate effects or prevalence may strongly impact optimality [16]. Regarding covariate optimisation, methods have been proposed to optimally select subjects to be included in a study with survival outcome based on a discrete covariate [17]. In case of discrete-time survival models, methods to compute the optimal allocation across different values of a continuous covariate [18], or between treatment groups and placebo groups [19] have been proposed. Nonetheless, these approaches do not extend to models including random effects to account for the inter-individual variability (IIV) due to the lack of analytical solution for the FIM. For discrete-time survival models including random effects, computation strategies based on grid search or particle swarm optimization (PSO) have been proposed to derive optimal designs [20]. Promising alternative for survival outcomes with random effects, and more generally, for non Gaussian continuous outcomes, may be FIM computation based on Monte-Carlo (MC) integration over observations combined with Markov Chains Monte-Carlo [21] (MCMC) or Adaptive Gaussian Quadrature [22] (AGQ) to integrate over random effects.

Thus, optimal design strategies for longitudinal continuous responses on the one hand and survival outcomes in the other hand have been addressed separately. To our knowledge, no work has targeted optimal design for joint models linking continuous NLMEM and time-to-event responses. Yet, the link between the longitudinal and survival processes implies that design decisions affecting one process inherently affect the other. This calls for dedicated optimal design methods capable of accounting simultaneously for the non-linear mixed-effects structure of the biomarker trajectory and the time-to-event component of the model. Our aim is therefore to develop such optimal design strategies and to illustrate their benefits for clinical study design. In this work, we extend Fisher Information Matrix–based design evaluation to joint models linking continuous non-linear mixed-effects models and time-to-event outcomes. We propose two computational strategies, MC-AGQ and MC-MCMC, that enable accurate evaluation of parameter uncertainty, power, and sample size across complex longitudinal–survival designs, including covariate optimisation. To our knowledge, this is the first general framework allowing FIM-based design evaluation and optimisation for such joint models.

The paper is organised as follows. Section 2 introduces the FIM computation for models linking continuous NLMEM and time-to-event responses. Section 3 describes a motivating joint tumour size–survival example based on the control arm of HORIZON III clinical study [23]. In section 4, the proposed methods are evaluated in regard to the influence of hyper-parameters and their performances in comparison with CTS on this tumor-size-survival joint model. Section 5 presents an extension of this example, illustrating the use of the MC-AGQ method to compare clinical trial designs and to optimise the discrete covariate distribution among the subjects to be included. Section 6 concludes with a discussion and perspectives for future research.

## 2 FIM computation for joint models

In this section, we first define population design and joint models, then derive the FIM expression, and finally present two numerical strategies for its computation.

### 2.1 Design

In this work, we focus on the design of the observations resulting from a joint model linking longitudinal continuous measurements and a single time-to-event outcome.

We denote Ξ = {*N*, (*ξ*_1_, …, *ξ*_*N*_), *p*_*Z*_} the planned population design for the data collection, where *N* is the number of subjects, the *ξ*_*i*_ are their elementary designs and *p*_*Z*_ is the covariate distribution in the target population. The elementary design 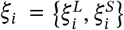 for a subject *i* contains all design information specified at the planning stage for both the longitudinal and survival processes. In particular, the longitudinal design 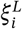 may include the planned number of longitudinal measurements *n*_*i*_, the corresponding measurement times 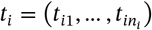, and the definition to compute a planned maximal follow-up duration 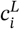. The survival design 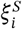 may include the planned follow-up duration 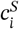, and time intervals for event assessment. In this work, we will only consider continuous-time observation for the event, so that 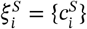. In many practical settings, 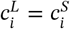 and corresponds to a time since the beginning of the study or a duration since patient inclusion.

However, due to the occurrence of the event process and censoring, the observed design may differ from the planned design. In particular, denoting *T*_*i*_ the time at which the event occurs for the *i*^*th*^ subject, longitudinal measurements are only observed up to time min 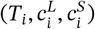, and follow-up is terminated after the event time *T*_*i*_ when 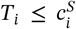. As a consequence, the effective number and timing of observed longitudinal measurements are random and subject-specific.

### 2.2 Joint Models

The observation vector writes 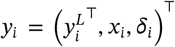, where 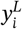 contains longitudinal measurements taken before min 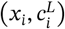 and where 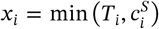 is the minimum between the event time *T*_*i*_ and the event censoring time 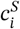 for this subject, and δ_*i*_, which equals 1 in case the event has been observed 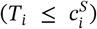 and 0 otherwise.

The vector of longitudinal continuous outcomes 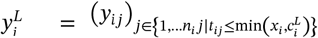 for subject *i* is modelled with a NLMEM:

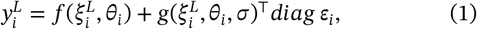

where *f* is the structural non-linear model, which depends on 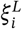 and *θ*_*i*_ = *u* (*μ, β*_*L*_, *z*_*i*_, *η*_*i*_), the individual parameter vector. The residual error is modelled through the function *g*, which additionally depends on some parameters *σ*, and on *ε*_*i*_ ~ 𝒩 0, *I*_*ni*_.

The individual parameters are a function of *μ*, the typical values, *β*_*L*_, some covariate effects, *z*_*i*_, the covariate vector and *η*_*i*_, the *d*-vector of individual random effects: *η*_*i*_ ~ 𝒩(0, Ω).

The time-to-event is modelled through the hazard function,

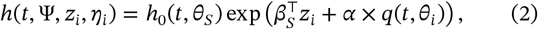

where *h*_0_(*t, θ*_*S*_), is the baseline hazard depending on some parameters gathered in *θ*_*S*_; *β*_*S*_ are some covariate effects; *α* is the link parameter and *q* is the link function which can depend on time and on the individual parameters *θ*_*i*_. It is common to have *q* = *f, q* = log *f* or 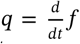 as link function; these choices correspond respectively to current value, log-current value, or slope of the longitudinal process.

The *P*-vector containing all the population parameters is denoted Ψ = (*μ, β*_*L*_, Ω, *σ, θ*_*S*_, *β*_*S*_, *α*).

### 2.3 FIM expression

For a population dessign Ξ, the FIM is obtained by aggregating the information matrices associated with each elementary designs:

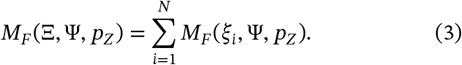

Given the distribution *p*_*Z*_ for the covariate vector, shared by the whole trial population, the FIM writes as an expectation with respect to this distribution [24]:

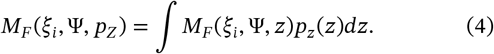

This integral can be approximated using Gauss-Legendre Quadrature and copula modelling for the covariate distribution [25]. In that case, the FIM is a weighted sum of FIM computed for each quadrature nodes combination.

Under quite general regularity conditions, the population FIM for an elementary design *ξ*_*i*_, given a covariate realisation *z*_*i*_ writes:

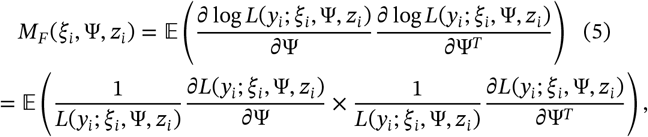

where *L* denotes the likelihood of the observations. The expectation in equation (5) is taken with respect to the joint distribution of the observations, 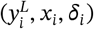 under the model. This means that by definition, the FIM accounts for the randomness of the observed design.

The likelihood of the observation is given by:

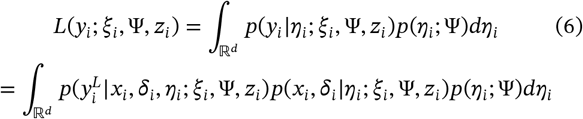

Accounting for censoring, we have that

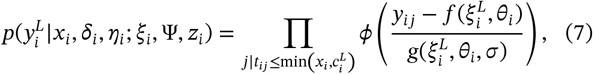

where *ϕ* denotes the density of the standard normal distribution. The survival contribution to the likelihood writes

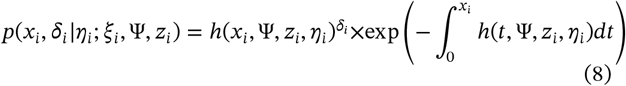

Expected SE on the parameters are computed as the square roots of the diagonal elements of the inverse of the FIM.

Designs can be compared based on the D-criterion [10],

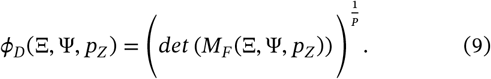

The larger *ϕ*_*D*_, the smaller the volume of the confidence ellipsoid for the parameter estimates, indicating higher precision.

### 2.4 FIM computation

In this work we consider two approaches which both use a Monte-Carlo approximation to compute the integral over the observations of equation (5):

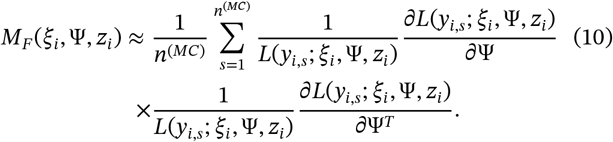

#### 2.4.1 MC-AGQ

In this first approach [22], a Gaussian quadrature is used to compute the integral over the random effects in the marginal likelihood and its derivatives:

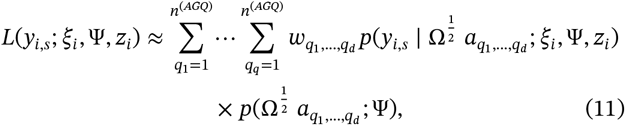

where 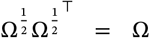 More precisely, an Adaptive Gaussian quadrature (AGQ) is performed, to take the shape of the integrand into account. Thus, nodes are centred around the mode of the joint distribution between the observations and the random effects,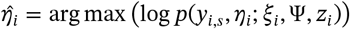, and rescaled to match the curvature,

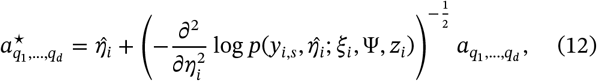

where 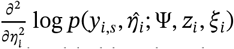 denotes the second derivative of the joint log-likelihood with respect to the random effects and evaluated at 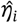. Weights are adapted accordingly:

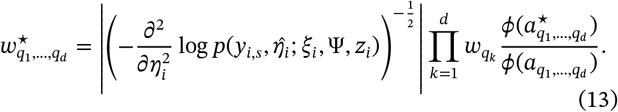

#### 2.4.2 MC-MCMC

The second approach [21] relies on a rewriting of 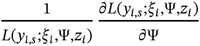 as a conditional expectation given *y*_*i,s*_:

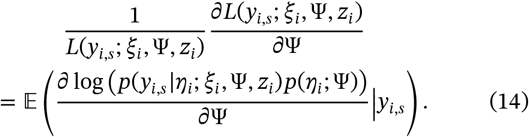

Finally, the MC approximation for the FIM writes:

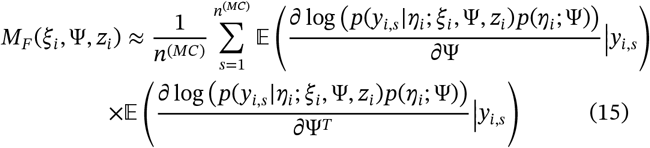

Therefore, two MCMC chains are needed to evaluate each of the conditional expectation:

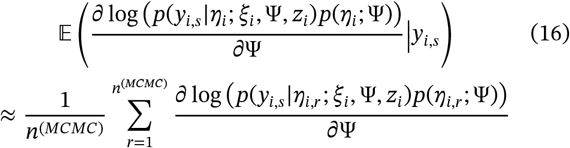

where *η*_*i,r*_, *r* = 1, … *n*^(*MCMC*)^ are drawn in the conditional pdf of *η* given *y*_*i,s*_.

#### 2.4.3 Monte-Carlo confidence interval

The proposed algorithms rely on stochastic numerical approximations, which induce variability in the estimated FIM. For both MC-AGQ and MC-MCMC, the variability due to the MC integration can be quantified to assess the numerical uncertainty associated with the design criteria using a bootstrap procedure. From the *n*^(*MC*)^ Monte-Carlo samples 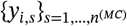, we generate *B* bootstrap resamples by sampling with replacement *n*^(*MC*)^ indices from {1, …, *n*^(*MC*)^}, denoted 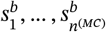. For each boot-strap sample 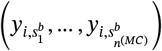 an approximation of the FIM is computed, 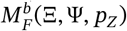, with its D-criterion. Empirical quantiles of the resulting distributions of the D-criterion and relative standard errors (RSE) are then used to derive uncertainty intervals reflecting the Monte-Carlo variability of the numerical FIM approximation.

## 3 Motivational example

Our example was inspired the control arm of HORIZON III clinical study [23], a clinical trial conducted in patients with metastatic colorectal cancer. In this control arm, patients were intended to be treated with FOLFOX6 and bevacizumab every 2 weeks.

### 3.1 Data

The control arm data were made publicly available on Project data sphere (https://data.projectdatasphere.org/projectdatasphere/html/content/78), and previously used in a methodological work on Bayesian inference on joint models [26]. Based on this publication, the considered dataset, which excluded patient with major deviation to the protocol, was composed of *N* = 593 subjects with 244 (41.15%) Females, and 87 (14.67%) subject with lactate dehydrogenase (*LDH*) higher than 1.5 times the upper limit of normal (ULN). The average Alkaline Phosphatase (ALP) was 165.34 U/L, with 10^*th*^ and 90^*th*^ being 64.41 and 322.33 U/L respectively. Summary metrics for the four groups obtained by combining the *Sex* and the *LDH* binary status are given in Table 1.

**TABLE 1.** Horizon data, covariate data summary and median survival timea cross the four discrete covariate combinations.

| SEX | LDH | N (%) | mean ALP (U/L) [P10, P90] | median survival time (years) |
| --- | --- | --- | --- | --- |
| Male | LDH ≤ 1.5ULN | 310 (52.28) | 146.89 [146.89, 146.89] | 2.14 |
|  | LDH > 1.5ULN | 39 ( 6.58) | 297.92 [297.92, 297.92] | 1.60 |
| Female | LDH ≤ 1.5ULN | 196 (33.05) | 136.76 [136.76, 136.76] | 2.13 |
|  | LDH > 1.5ULN | 48 ( 8.09) | 293.42 [293.42, 293.42] | 1.31 |

The tumour size was assessed by the sum of longest diameter (SLD) of target lesions. The baseline measurement was taken four weeks before treatment initiation. Then, SLD was measured while on-treatment, every 8-weeks until week 24, and subsequently every 12 weeks until disease progression or death. Among the patients considered, the number of measurements ranged between 2 and 14 but most of the subjects had only 6 or 5 SLD measurements (respectively 25.46 and 21.24%). Longitudinal SLD measurements are shown on Figure 1 for each of the 4 discrete covariates groups. Overall, death was observed for 195 subjects (32.88%). The median survival for each of the 4 discrete covariates groups are given in Table 1 and the Kaplan-Meier curves of survival are shown on Figure 1.

**FIGURE 1.**
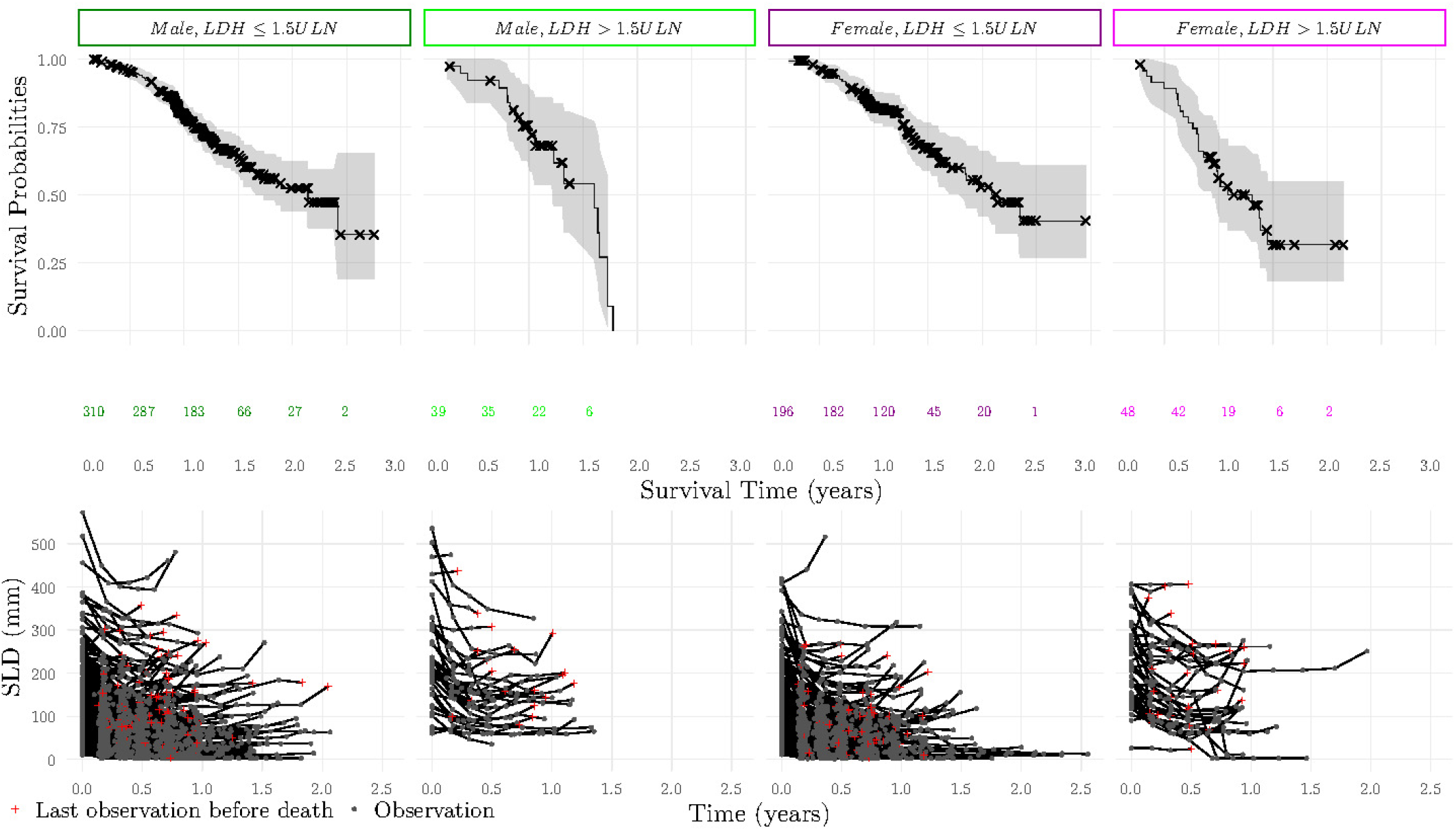
Horizon data, Kaplan-Meier curves of survival and longitudinal measurements of sum of lesion diameter (SLD) across the four discrete covariate combinations.

### 3.2 Joint Model

The joint model describing the SLD and the survival was taken from [26]. SLD dynamics followed a Stein-Fojo model and was modelled with a proportional error model, 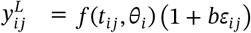 and

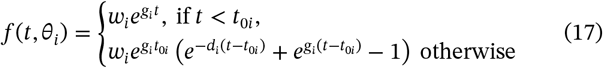

where *t*_0*i*_ indicates the time when the treatment was initiated. Individual parameters followed independent log-normal distributions:

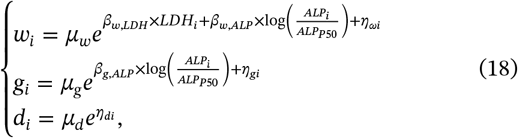

with 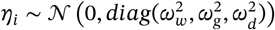.

Survival was modelled by a Weibull hazard with the individual tumour growth rate *g*_*i*_ as link function:

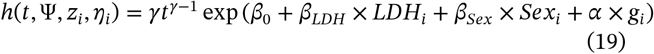

As in [26] two binary covariates, *Sex* and *LDH* were included in the survival sub-model (equation (19)). Additional covariates relationships were included as additive effects on the logscale on the longitudinal sub-model by running the COSSAC algorithm [27] and keeping only the relevant relationships. Continuous covariates were normalised by the median, *z*_*P*50_, of the entire dataset and log-transformed, 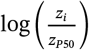 Relevance was defined based on the 90% confidence interval of the ratio, with the relevance area set to [0.80; 1.25]. The final model included *LDH* effect on baseline SLD, and Alkaline Phosphatase (ALP) on baseline SLD and tumour growth rate (equation (18)).

Parameters used in the following of our study were estimated using the SAEM algorithm implemented in Monolix 2024R1 [28] (Table 2). 10 chains were used in the Metropolis-Hastings algorithm for individual parameters estimation at each iteration. Parameters were initialised to the posterior mean obtained with Bayesian inference published in [26].

**TABLE 2.**
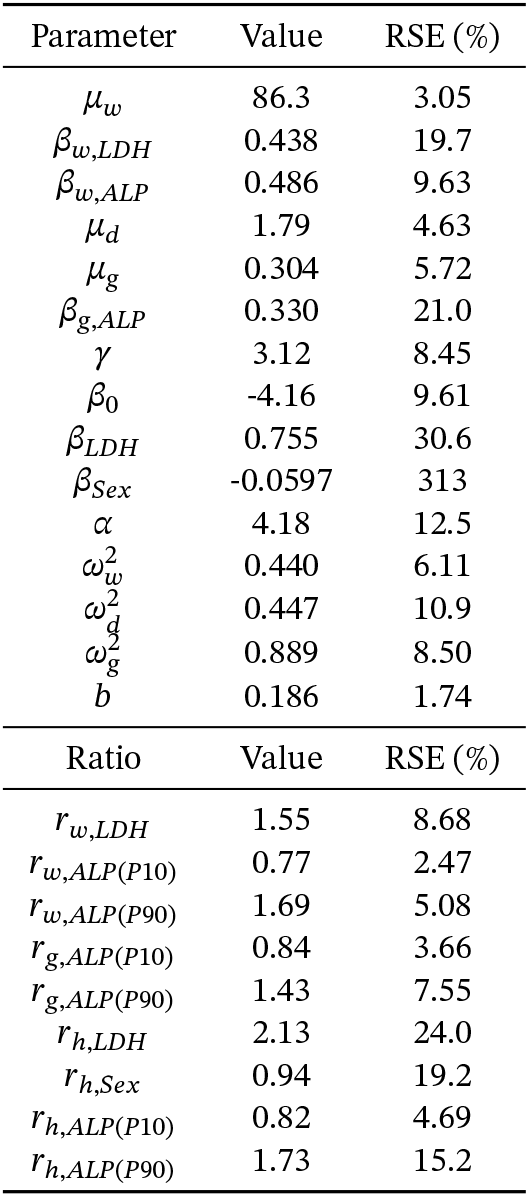
Parameter values and derived covariate ratios, estimated using the SAEM algorithm on data from the Horizon study and their relative standard errors (RSE).

### 3.3 Ratio computation for covariate assessment

The magnitude of covariate effects was assessed by computing the ratio of change in primary parameters associated with a given covariate value and relative to a reference value. The uncertainty on this ratio can be derived from the SE on the covariate effect parameter, and relevance and non-relevance tests can be performed [24].

For discrete covariates, ratios of their effects on longitudinal parameters and on the hazard were computed as 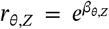. For continuous covariates, ratios were computed for the 10^*th*^ and 90^*th*^ percentiles of the covariate distribution, denoted *z*_*P*10_ and *z*_*P*90_, and relatively to its median: 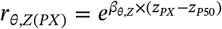. Of note, if a continuous covariate *Z* has an effect on the tumour growth rate which is the link function, the ratio of the effect of *Z* on the hazard can also be computed as:

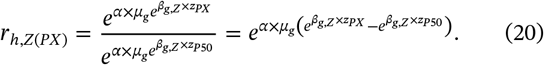

Because the covariates were normalised by the median and log-transformed, *z*_*P*50_ = 0, and therefore

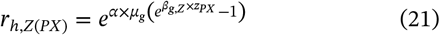

The Delta method can be used to derive the uncertainty on this ratio, and the resulting normal approximation can be used to performed statistical test on the relevance/non-relevance of the relationship. The ratio derivatives are given by:

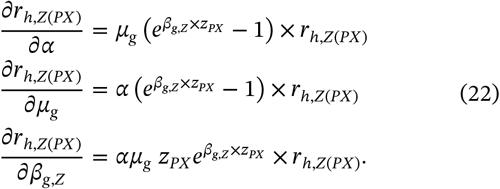

Estimated ratios on the Horizon database are given in Table 2. With respect to the equivalence interval [0.80; 1.25], the effect of *LDH* on the SLD baseline *w* (1.55) and on the survival hazard *h* (2.13) have to be shown relevance, as long as the effect of the 90^*th*^ *ALP* percentile on the SLD baseline (1.69), on the tumour growth rate (1.43) and on *h* (1.73). These ratios are consistent with literature, as both high *LDH* levels and elevated *ALP* were shown to be associated with shorter overall survival in colorectal cancer patients [29, 30]. The effect of Sex on *h* has to be shown non-relevant (0.94). Of note, the effects on the 10^*th*^ *ALP* percentile are very close to the equivalence area borer, so unlikely to be shown relevant or non-relevant.

## 4 Evaluation of the algorithms

The AGQ and MCMC FIM computation methods described in Section 2 were implemented in R, editing respectively a previous implementation of the AGQ approach, available at https://github.com/saemixdevelopment/saemixextension/tree/master/fimAGQ and the R package *MIXFIM* [31]. The resulting implementation was then evaluated the tumour-growth–survival model described in Section 3 regarding the influence of algorithm hyper-parameters and the performance of the FIM approximation obtained in comparison with results from CTS.

### 4.1 Methods

#### 4.1.1 Design

We considered a design including *N* = 250 subjects with the start of the study at the initiation of treatment (i.e. *t*_0_ = 0). SLD were measured at baseline and then with 8-week intervals until week 24 and subsequently every 12 weeks until death, up to 3 years. This sampling is hereafter referred to as rich sampling. In that case we have 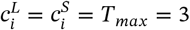 years for all the subjects. To better reflect real life condition, we also considered a design in which patients with disease progression according to Response Evaluation Criteria in Solid Tumors (RECIST) version 1.0 [32] may be offered a second line therapy and their SLD measurements are stopped but information for survival is kept. Thus, 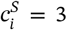 years for all the subjects, but 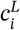 depends on the trajectory of the SLD. Results for this scenario are presented in Appendix B.

#### 4.1.2 FIM computation

##### 4.1.2.1 Covariate distribution

To integrate the FIM over the distribution of the covariates included in the model, this distribution first need to be characterised. We therefore fitted a kernel density estimate for the ALP on the four subsets of the Horizon database corresponding to each discrete covariate combination, using the R package *rvinecopulib*0.7.3.1.0 [33]. Goodness-of-fit was evaluated using the relative error on summary statistics for continuous covariates, as well as visual predictive checks [34]. Diagnostics were performed using 100 covariate datasets simulated from the copula, each containing the same number of subjects as the corresponding Horizon subset.

##### 4.1.2.2 Hyper-parameters tuning

Integral over covariate distribution was approximated with Gauss-Legendre quadrature. To determine the number of nodes required to achieve acceptable accuracy, we computed the FIM with *n*^(*GLQ*)^ = 1, 3, 5 and 7, and chose *n*^(*GLQ*)^ to reach a relative convergence in D-criterion below 2.5% between to tested number of nodes.

To select the number of MC sample needed, computations were performed up to *n*^(*MC*)^ = 3000. The final choice ensured that the 95% bootstrap confidence interval on D-criterion has a relative width below 5%.

For MC-AGQ and MC-MCMC, the number of quadrature nodes and MCMC samples were chosen respectively from *n*^(*AGQ*)^ ∈ {1, 3, 5, 7} and *n*^(*MCMC*)^ ∈ {100, 200} after 500 warm-up samples, in order to reach a relative convergence below 2.5% in D-criterion.

#### 4.1.3 Comparison with CTS

As a benchmark and target reference, CTS were performed by simulating 200 datasets in R to ensure stable empirical variances estimates, with covariate vectors sampled with replacement from the Horizon database. Parameter estimation was performed using the SAEM algorithm implemented in Monolix 2024R1, using 10 chains and initial points set at the simulation values.

The empirical D-criterion was computed from the empirical variance-covariance matrix and compared with the D-criteria obtained from the FIM computed using MC-AGQ and MC-MCMC. Expected SE were calculated as the square roots of the diagonal elements of the inverse FIM computed with MC-AGQ and MCMCMC, while empirical SE were computed as the standard deviation of the parameter estimates across the simulated datasets. For easier comparison, relative standard errors (RSE) were reported.

### 4.2 Results

#### 4.2.1 FIM computation

*ALP* distribution was correctly estimated for each of the four covariate combinations as shown in Appendix A, Figures A1 and A2.

Figure 2 **A** (a) shows the D-criterion as a function of the number of Monte-Carlo samples (*n*^(*MC*)^), for increasing number of quadrature nodes in covariate integration (*n*^(*GLQ*)^) and increasing number of quadrature nodes in random effects integration (*n*^(*AGQ*)^) when computing the FIM with MC-AGQ. D-criterion was quite stabilised from *n*^(*MC*)^ = 1000, and at least *n*^(*AGQ*)^ = 5 was needed as there was no overlap in confidence intervals between *n*^(*AGQ*)^ = 3 and *n*^(*AGQ*)^ = 5, no matter *n*^(*GLQ*)^. With all the explored *n*^(*GLQ*)^ and *n*^(*AGQ*)^, the 95% bootstrap CI on *ϕ*_*D*_ was below 5% from *n*^(*MC*)^ = 1000 and higher (Figure 2 **A** (b)). This results suggested that *n*^(*MC*)^ = 1000 was enough to reach the desired convergence.

**FIGURE 2.**
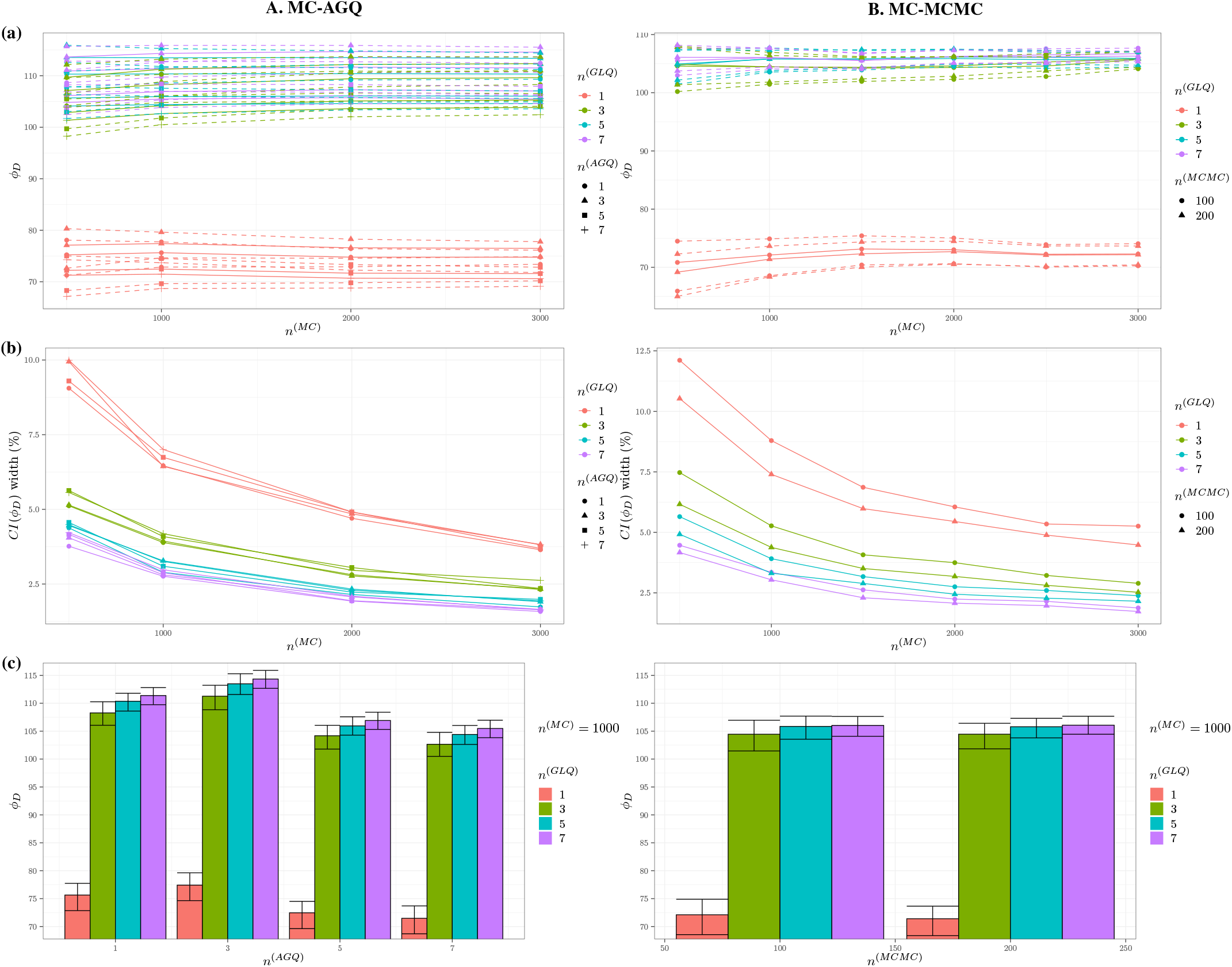
Influence of the hyper-parameters in Fisher Information Matrix computation using Monte-Carlo and Adaptive Gaussian Quadrature (MC-AGQ) on the left, and Monte-Carlo and Markov-Chain Monte-Carlo (MC-MCMC) on the right **(a)** D-criterion (*ϕ*_*D*_) with its 95% confidence interval and **(b)** bootstrap confidence interval on *ϕ*_*D*_ relative width, as a function of the number of Monte-Carlo samples (*n*^(*MC*)^), for increasing number of quadrature nodes in covariate integration (*n*^(*GLQ*)^) and respectively, increasing number of quadrature nodes in random effects integration (*n*^(*AGQ*)^) for AGQ and increasing number of MCMC samples in random effects integration (*n*^(*MCMC*)^) for MCMC **(c)** D-criterion (*ϕ*_*D*_) with its 95% confidence interval with *n*^(*MC*)^ = 1000 and increasing number of quadrature nodes in covariate integration (*n*^(*GLQ*)^) and respectively increasing *n*^(*AGQ*)^ and *n*^(*MCMC*)^.

Figure 2 **A** (c) shows, with *n*^(*MC*)^ = 1000, a drop in D-criterion between *n*^(*AGQ*)^ = 3 and *n*^(*AGQ*)^ = 5, and then almost same results between *n*^(*AGQ*)^ = 5 and *n*^(*AGQ*)^ = 7, suggesting to keep *n*^(*AGQ*)^ = 5. With *n*^(*MC*)^ = 1000, with *n*^(*AGQ*)^ = 5, CI overlapped and increasing the number of nodes *n*^(*GLQ*)^ in the covariate integration from 5 to 7 changed *ϕ*_*D*_ by less than 1%, therefore *n*^(*GLQ*)^ = 5 was chosen With *n*^(*MC*)^ = 1000 and *n*^(*GLQ*)^ = 5, the D-criterion was respectively 105.9 [104.2, 107.5] and 104.4 [102.7, 106.0] when using either *n*^(*GLQ*)^ = 5 and *n*^(*GLQ*)^ = 7, with respective runtimes of 1.1 hour and 3.2 hours for FIM computation (Appendix A, Table A1). The relative difference in *ϕ*_*D*_ between these two *n*^(*GLQ*)^ was 1.45%, therefore below 2.5 %.

Thus the hyper-parameters *n*^(*MC*)^ = 1000, *n*^(*GLQ*)^ = 5, *n*^(*AGQ*)^ = 5 were selected, corresponding to a run time of 1.1 hour.

Figure 2 **B** (a) shows the D-criterion as a function of the number of Monte-Carlo samples (*n*^(*MC*)^), for increasing number of quadrature nodes in covariate integration (*n*^(*GLQ*)^) and increasing number of MCMC samples in random effects integration (*n*^(*MCMC*)^) when computing the FIM with MC-MCMC. D-criterion was quite stabilised from *n*^(*MC*)^ = 1000, and at least *n*^(*GLQ*)^ = 3 was needed as there was no overlap in confidence intervals between *n*^(*GLQ*)^ = 3, centred around 70 and the other values that are centred around 105. The results for *n*^(*MCMC*)^ = 100 and *n*^(*MCMC*)^ = 200 were very close, suggesting that *n*^(*MCMC*)^ = 100 was enough. For the bootstrap CI on *ϕ*_*D*_ to be below 5% with *n*^(*MC*)^ = 1000, at least 3 quadrature nodes or more in covariate integration are needed with *n*^(*MCMC*)^ = 200 but 5 or more with *n*^(*MCMC*)^ = 100 (Figure 2 **B** (b)). With *n*^(*MCMC*)^ = 100, for equal *n*^(*GLQ*)^, D-criterion 95% CI greatly overlap between *n*^(*GLQ*)^ = 3, 5 and 9 (Figure 2 **B** (c)), therefore *n*^(*GLQ*)^ = 5 was enough to have less than 2.5% difference between two tested number of nodes. With *n*^(*MC*)^ = 1000 and *n*^(*GLQ*)^ = 5, there was almost no difference in *ϕ*_*D*_ between *n*^(*MCMC*)^ = 100 and *n*^(*MCMC*)^ = 200 (respectively 105.8 [103.6, 107.7] and 105.8 [103.8, 107.3]), therefore because the computation time was shorter *n*^(*MCMC*)^ = 100 was kept (3.0 h vs 3.6 h, Appendix A, Table A2).

Finally, the hyper-parameters *n*^(*MC*)^ = 1000, *n*^(*GLQ*)^ = 5, *n*^(*AGQ*)^ = 5 and *n*^(*MC*)^ = 1000, *n*^(*GLQ*)^ = 5, *n*^(*MCMC*)^ = 100 were selected for respectively MC-AGQ and MC-MCMC FIM computation and are hereafter referred to as tuned MC-AGQ and MC-MCMC. They respectively ran in 1.1 hour and 3.0 hours.

In Appendix B are given the D-criterion convergence depending on the hyper-parameters both for both MC-AGQ and MC-MCMC (Figure B6) in the scenario were SLD was censor in case of disease progression. Hyper-parameters selected on the scenario without censor are still valid.

All the R codes needed to run the FIM computation with MC-AGQ and MC-MCMC with the selected hyper-parameters are available online at: https://doi.org/10.5281/zenodo.20430052.

#### 4.2.2 Comparison with CTS

Simulated datasets contained in average 2408 (sd = 59.6) SLD observations and an example of simulated dataset is shown in Appendix A, Figure A3. The total estimation run time for the CTS with 200 datasets was about 100 h, therefore much longer than the tuned MC-AGQ and MC-MCMC which respectively ran in 1.1 and 3.0 h in the scenario without SLD censoring.

The empirical D-criterion obtained with CTS was 109.96 [109.92; 121.15], therefore close to FIM predictions, respectively 105.9 [104.2, 107.5] using tuned MC-AGQ and 105.8 [103.6, 107.7] using tuned MC-MCMC.

Figure 3 shows an overall good consistency between the predicted RSE obtained through CTS with 200 datasets and those obtained via FIM computation using tuned MC-AGQ and MC-MCMC. Of note, the uncertainty associated with the effect of *LDH* on survival appears to be under-predicted by the FIM. This is not unexpected, as the FIM provides a lower bound on the uncertainty, which is reached only in asymptotic settings. In the present case, subjects with high *LDH* represent less than 15% of the trial population of size *N* = 250, corresponding to approximately 37 individuals.

**FIGURE 3.**
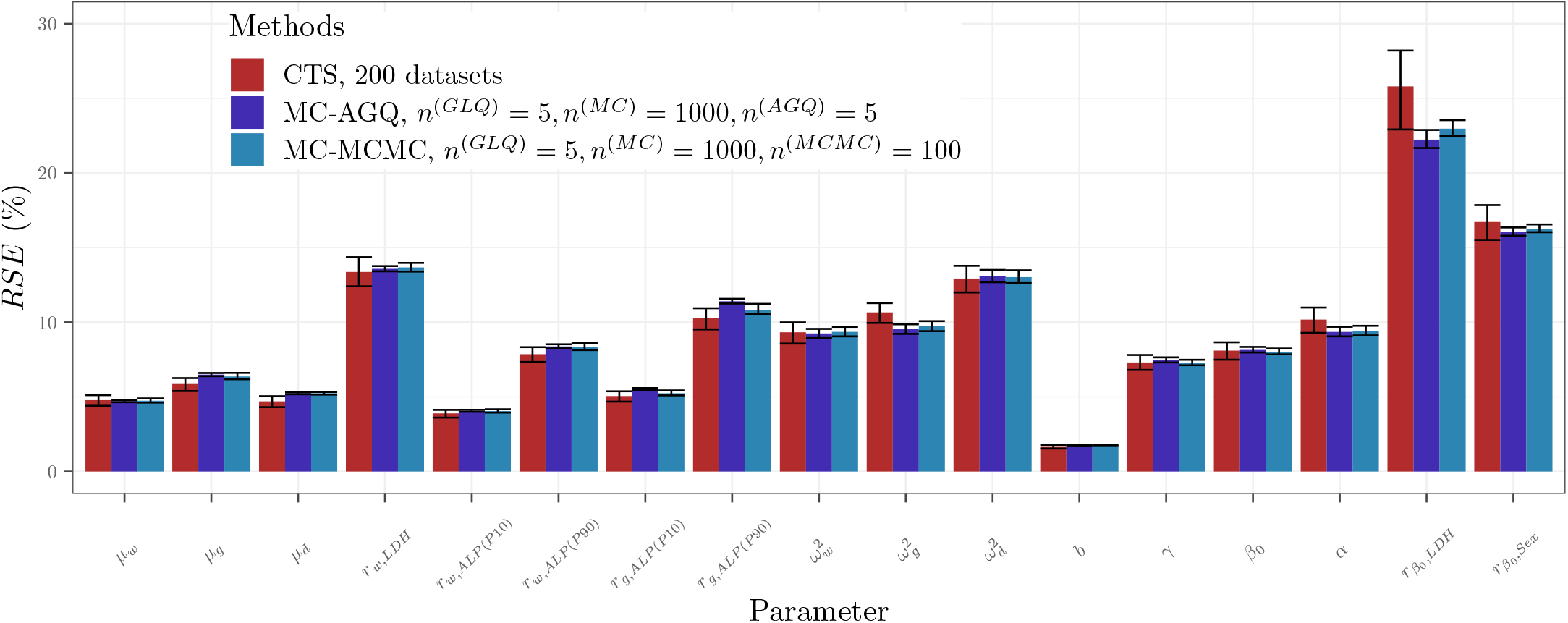
Predicted Relative Standard Errors (RSE) obtained through Clinical trial Simulations (CTS) with 200 datasets, FIM computation using Monte-Carlo and Adaptive Gaussian Quadrature (MC-AGQ) with *n*^(*MC*)^ = 1000, *n*^(*GLQ*)^ = 5, *n*^(*AGQ*)^ = 5, and FIM computation using Monte-Carlo and Markov-Chain Monte-Carlo (MC-MCMC) with *n*^(*MC*)^ = 1000, *n*^(*GLQ*)^ = 5, *n*^(*MCMC*)^ = 100

In the scenario with censored SLD measurements, simulated datasets contained in average 1311 (sd = 38.9) SLD observations and an example of simulated dataset is shown in Appendix B, Figure B5. In this scenario, RSE were higher (Figure B7), while still showing very good agreement between the empirical RSE obtained from CTS and those predicted by the FIM computed using both tuned MC-AGQ and MC-MCMC for longitudinal parameters. However, uncertainty was consistently under-predicted for all survival parameters. The fact that MC-AGQ and MC-MCMC yielded very close results suggests that the FIM was accurately approximated. Therefore, this discrepancy is more likely attributable to a known limitation of the FIM, as it is only an asymptotic approximations of parameter uncertainty and in this scenario much less SLD data are available an asymptotic conditions may not be reached.

## 5 Application of the algorithm for design evaluation and optimisation

The section 4 shown the accuracy of both MC-AGQ and MC-MCMC methods to efficiently predict uncertainty on model parameters compared to CTS. This section illustrates how this can be leveraged to compare various designs and optimise covariate allocation before conducting a clinical study. Because MC-AGQ was faster, only this method was kept for this section.

### 5.1 Methods

The influence of the design was studied by varying the follow-up duration, *T*_*max*_, between 1, 2 and 3 years, as well as by modifying the sampling richness and the sample size. We defined the rich sampling as the one used in the previous section, consisting of a baseline SLD measurement followed by assessments every 8 weeks up to week 24 and then every 12 weeks until death. For a 3-year follow-up, it corresponds to observations at weeks 0, 8, 16, 24, 36, 48, 60, 72, 84, 96, 108, 120, 132, 144 and 156. The sparse sampling was obtained by keeping one observation out of two from the rich design.

The optimal distribution between the four Sex × *LDH* combinations was computed using the Projected Gradient Descent algorithm, using the D-criterion as objective function [25].

FIM was computed using MC-AGQ with *n*^(*GLQ*)^ = 5, *n*^(*MC*)^ = 1000 and *n*^(*AGQ*)^ = 5 as selected in section 4.

Designs and covariate distributions were compared based on the associated D-criterion; RSE on all the parameters and especially on *α*, the link parameter; power of relevance/non-relevance tests for the covariate effects and *NSN* to reach 80% power in these tests.

### 5.2 Results

As expected, the longer the follow-up duration and the richer the sampling, the more informative were the designs (Figure 4 (a)). Surprisingly, in our example longitudinal-parameter uncertainty was barely affected by follow-up duration or sampling richness (Figure 5). Uncertainty on survival-parameters decreased substantially from 1-year to 2-year follow-up, and 1-year follow-up was too short to identify discrete covariate effects, with RSE between 40 and 70%. Interestingly, although uncertainty in the link parameter decreased with longer follow-up, richer sampling, and higher sample size, it was very similar between a 2-year follow-up with rich sampling and a 3-year follow-up with sparse sampling (Figure 4 (b)). For instance the *NSN* to reach below 15% RSE on *α* was 120 [110, 130] with 2-year follow-up with rich sampling and 122 [114, 131] 3-year follow-up but sparse sampling.

**FIGURE 4.**
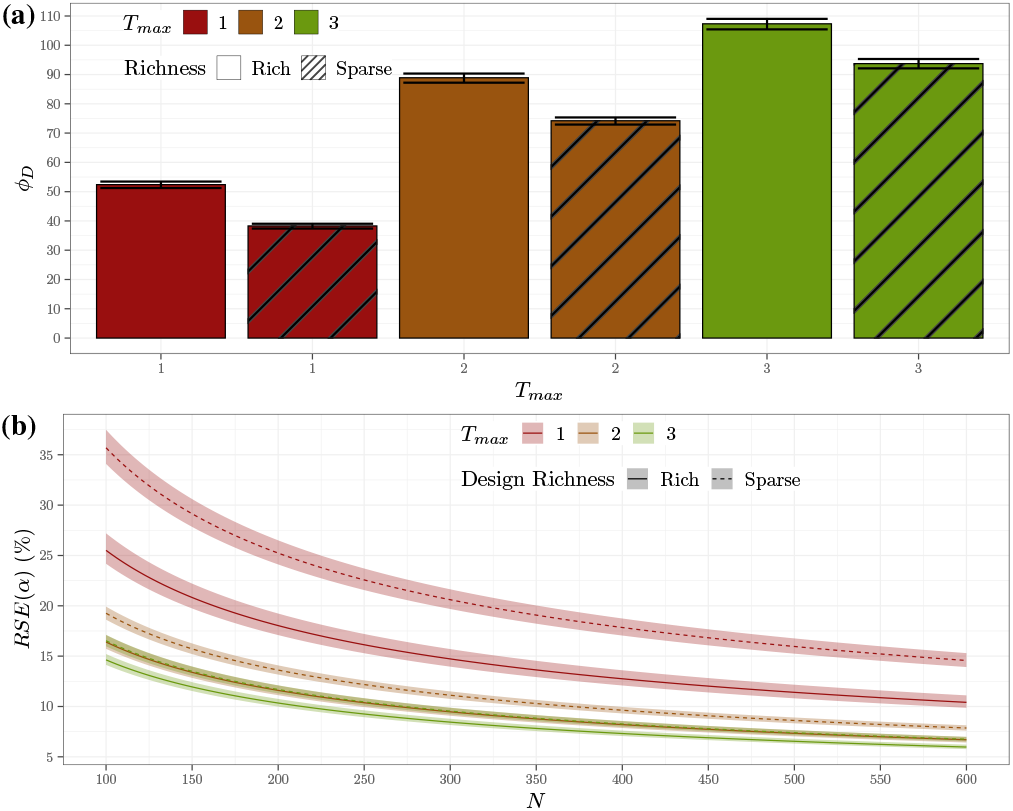
Comparison of designs based on Fisher Information Matrix computation using Monte-Carlo and Adaptive Gaussian Quadrature (MC-AGQ) with *n*^(*GLQ*)^ = 5, *n*^(*MC*)^ = 1000 and *n*^(*AGQ*)^ = 5 **(a)** D-criterion (*ϕ*_*D*_) with its 95% confidence interval **(b)** Relative Standard Errors (RSE) on *α*, the link parameter as a function of the sample size *N*.

**FIGURE 5.**
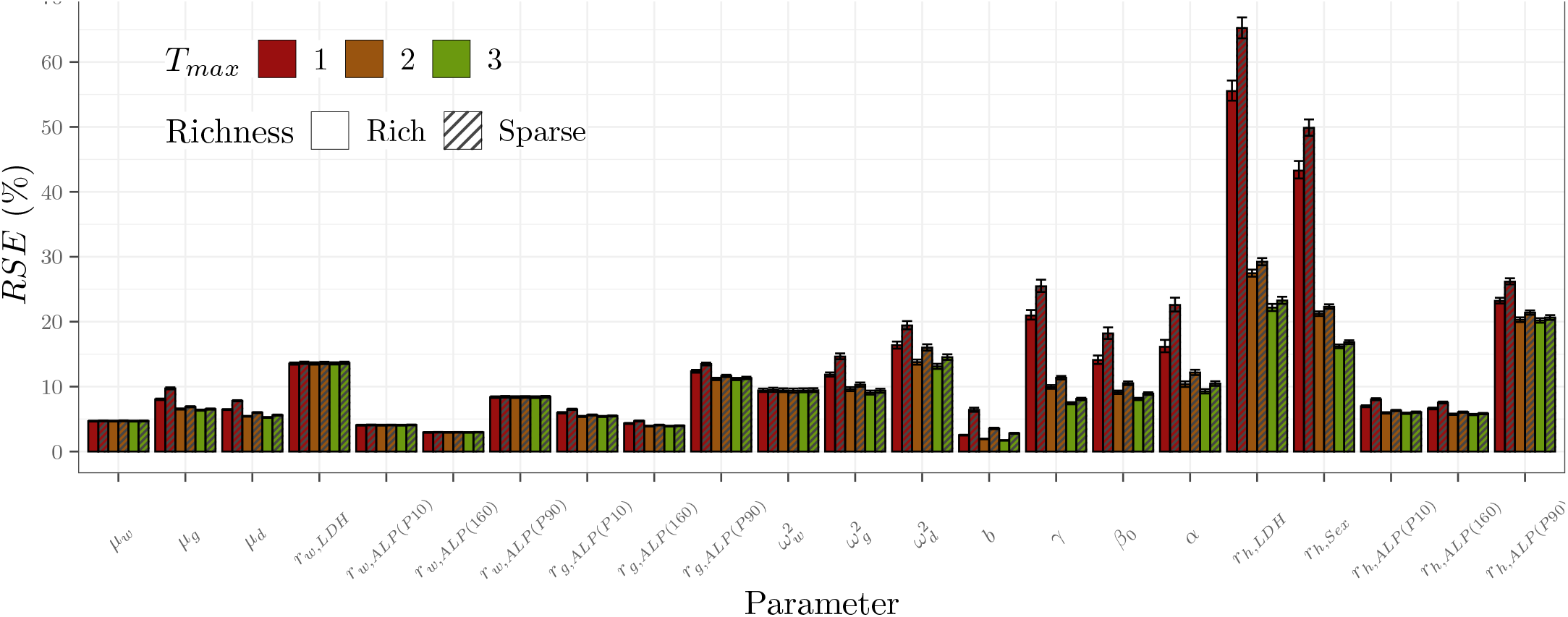
Comparison of Relative Standard Errors (RSE) for the various designs, based on Fisher Information Matrix computation using Monte-Carlo and Adaptive Gaussian Quadrature (MC-AGQ) with *n*^(*GLQ*)^ = 5, *n*^(*MC*)^ = 1000 and *n*^(*AGQ*)^ = 5.

Covariate optimisation always increased the informativeness of the design (Figure 6 (a)). The optimal distribution between the four discrete combinations was highly stable across designs and consistently required at least 45% of subjects with high *LDH* (Figure 6 (b)). Figure A4 in Appendix A shows that optimal covariate distribution always decreased the uncertainty on the covariate effects and therefore the width of the 90% CI on the ratios.

**FIGURE 6.**
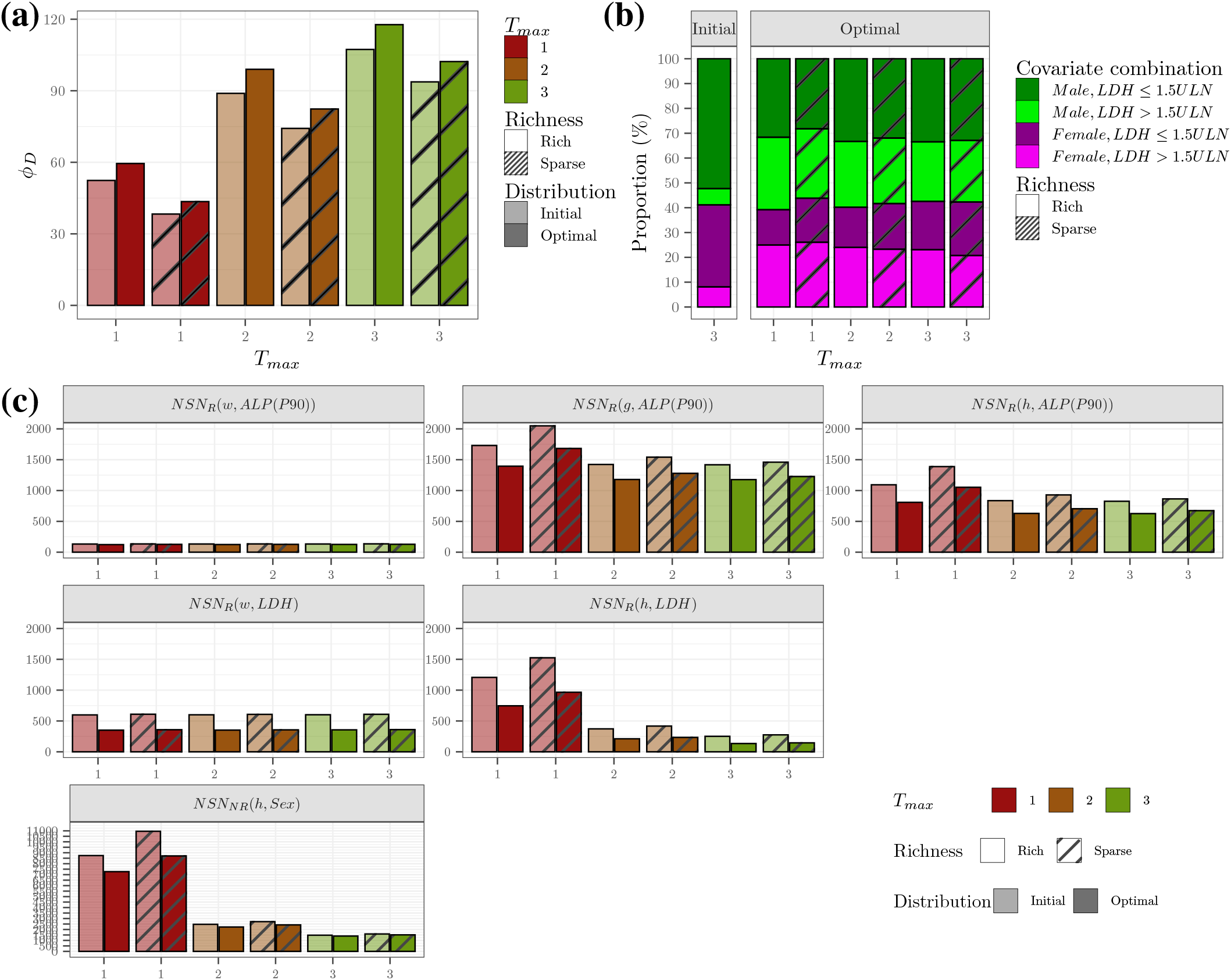
Covariate optimisation for various designs based on Fisher Information Matrix computation using Monte-Carlo and Adaptive Gaussian Quadrature (MC-AGQ) with *n*^(*GLQ*)^ = 5, *n*^(*MC*)^ = 1000 and *n*^(*AGQ*)^ = 5 **(a)** D-criterion (*ϕ*_*D*_) with its 95% confidence interval, **(b)** Optimal covariate distributions, **(c)** Forest plot, **(d)** Power of relevance and non-relevance tests on covariate relationships with [0.80; 1.25] as equivalence area, for the various designs and both initial and optimal covariate distributions.

Consequently, the optimal distribution always decreased the *NSN* to detect with 80% power *LDH* relevance on both baseline SLD and survival, as well as the *NSN* to detect the effect of the 90^*th*^ ALP percentile on the hazard ((Figure 6 (c)). Moreover, implementing the optimal distribution in a sparse 2-year follow-up design yielded smaller *NSN* than a rich 3-year follow-up design with the initial distribution. For instance, *NSN*_*R*_ (*h, LDH*) was 251 for the initial distribution with rich 3-year follow-up, while it was 233 with a 2-year follow-up design. Even more impressive, *NSN*_*R*_ (*h, ALP*(*P*90)) was 826 for the initial distribution with rich 3-year follow-up, while it dropped to 705 with a sparse 2-year follow-up design, corresponding to a decrease of almost 15%. In contrast, none of the investigated designs allowed to conclude on the non-relevance of *Sex* on the survival hazard for *N* = 250 and *NSN* to reach 80 % remained very high, ranging from 1402 for the optimal covariate distribution with a rich 3-year follow-up up to 10960 for a sparse 1-year follow-up.

## 6 Discussion

In this work two new computation approaches for FIM in joint models were introduced, relying on numerical approximation of the integral over the observations through MC and of the integral over random effects using either AGQ or MCMC.

The evaluation of the proposed algorithms was carried out through a comparison with CTS on a tumor-growth-survival joint model based on the control arm of the Horizon III study, for which a longitudinal covariate model was developed. The joint model thus included continuous and discrete covariates in the longitudinal sub-model and discrete covariates in the survival sub-model. Of note, the link function was the individual tumor growth rate, incorporating a continuous covariate effect of *ALP*. Therefore this joint modelling enabled to derive the indirect *ALP* impact on survival despite its absence from the survival sub-model. The proposed methodology further enabled the computation of the statistical power to detect the relevance of this effect on survival. In this work, the relevance region was defined as [0.8, 1.25]. However, this range should be adapted to the clinical context and determined in consultation with subject-matter experts.

Both MC-AGQ and MC-MCMC approaches were shown to be accurate and substantially faster than CTS. The MC-AGQ computation was the fastest and was therefore chosen to be applied on an example comparing multiple candidate designs. This investigation highlighted that similar information levels can be achieved either by extending the follow-up duration with sparser longitudinal sampling or by shortening the follow-up while increasing sampling richness. These results emphasize the trade-off between follow-up duration and sampling richness and illustrate how the proposed methodology can support informed design decisions. In addition, a covariate optimization framework was implemented using the MC-AGQ FIM computation and showed the influence of the covariate distribution on the power to detect the covariate effects on the survival hazard. More specifically, we have shown that, in our example, implementing the optimal covariate distribution makes it possible to achieve the same statistical power for detecting the clinical relevance of a prognostic covariate with a shorter follow-up duration, a smaller number of observations and a smaller sample size.

Our framework enables exhaustive design evaluation and optimisation, and provides a quantitative assessment of the *NSN* to achieve targeted levels of parameter uncertainty or statistical power for covariate testing. Although full design optimisation was not performed here, the availability of efficient FIM computation makes it straightforward to optimise sampling times or arm allocation using existing algorithms (see for instance [35, 36]). It should be noted, however, that the FIM relies on asymptotic approximations of parameter uncertainty, and its performance may therefore be limited for small sample sizes or sparse settings. This issue was encountered here in the scenario where SLD data were censored after disease progression; future work could explore in greater detail what sample size is required to achieve asymptotic conditions in various contexts, depending on the IIV, the strength of the link and the complexity of the model.

In our example, the link function was an individual parameter from the longitudinal sub-model, but both MC-AGQ and MC-MCMC computations can be applied with time-dependent link functions, such as the current value of the longitudinal process or a function of it. In such cases, computation times are expected to increase, as temporal integrals must be evaluated in the survival contribution to the likelihood. In addition, the method can be extended to context of competitive risk with multiple survival outcomes [6]. Despite this additional computational burden, the proposed framework remains a promising and flexible tool to assess the impact of design choices, especially across a range of link-parameter values. Indeed here we considered only local design, where parameters and models are assumed to be known, but this assumption is questionable, especially regarding the form of the link function and the link parameter value. Therefore, our FIM computation for joint model could be plugged in robust design frameworks already introduced for NLMEM context [37, 38, 39, 40, 41, 42].

A known limitation of the present case study is that only three random effects were considered. Due to the combinatorial explosion inherent to quadrature-based methods, AGQ may become impractical as the number of random effects increases. In such scenarios, MCMC-based approaches may constitute a more scalable alternative [21], although a systematic comparison of their respective performance in higher-dimensional settings should be further investigated. Similar limitations may arise when considering multiple continuous covariates. Of note, calibration of the numerical integrations was performed empirically and additional sensitivity analyses could be conducted to assess the influence of hyper-parameters on design decisions regarding sample size, sampling times and covariate distribution. Nevertheless, these differences are expected to remain acceptable for a first design evaluation and optimisation step, recalling that a final CTS step with the selected design is anyway recommended to validate the asymptotic approximation provided by the FIM. Furthermore, optimal study designs and optimal distributions of covariates must always be agreed upon in consultation with clinicians and practitioners to ensure both compliance with ethical principles and suitability for real-life conditions.

To conclude, we introduced a fast and accurate method for evaluating and optimising study designs based on a joint model of continuous and time-to-event outcomes. This approach efficiently quantifies the impact of design choices on parameter uncertainty and statistical power.

## Conflicts of Interest

The authors declare no conflicts of interest.

## Supporting Information

## APPENDIX

## A Appendix 1: Complementary results

**TABLE A1.** D-criterion (*ϕ*_*D*_) and its 95% bootsrap confidence interval and runtime for Fisher Information Matrix computation using Monte-Carlo and Adaptive Gaussian Quadrature (MC-AGQ) with *n*^(*MC*)^ = 1000 and *n*^(*GLQ*)^ = 5.

| $n^{(MC)}$ | $n^{(GLQ)}$ | $n^{(AGQ)}$ | $\phi_D$ [95% CI] | runtime (h) |
| --- | --- | --- | --- | --- |
| 1000 | 1 | 1 | 75.6 [ 72.9, 77.7] | 0.0 |
| 1000 | 3 | 1 | 108.3 [106.1, 110.3] | 0.1 |
| 1000 | 5 | 1 | 110.4 [108.6, 111.8] | 0.2 |
| 1000 | 7 | 1 | 111.4 [109.7, 112.8] | 0.2 |
| 1000 | 1 | 3 | 77.4 [ 74.6, 79.6] | 0.1 |
| 1000 | 3 | 3 | 111.3 [108.8, 113.2] | 0.2 |
| 1000 | 5 | 3 | 113.5 [111.6, 115.3] | 0.3 |
| 1000 | 7 | 3 | 114.3 [112.7, 115.9] | 0.4 |
| 1000 | 1 | 5 | 72.5 [ 69.6, 74.5] | 0.3 |
| 1000 | 3 | 5 | 104.2 [101.8, 106.0] | 0.7 |
| 1000 | 5 | 5 | 105.9 [104.3, 107.6] | 1.1 |
| 1000 | 7 | 5 | 106.9 [105.3, 108.4] | 1.5 |
| 1000 | 1 | 7 | 71.5 [ 68.7, 73.7] | 0.9 |
| 1000 | 3 | 7 | 102.7 [100.5, 104.8] | 1.7 |
| 1000 | 5 | 7 | 104.4 [102.6, 106.0] | 3.2 |
| 1000 | 7 | 7 | 105.5 [103.8, 107.0] | 5.0 |

**FIGURE A1.**
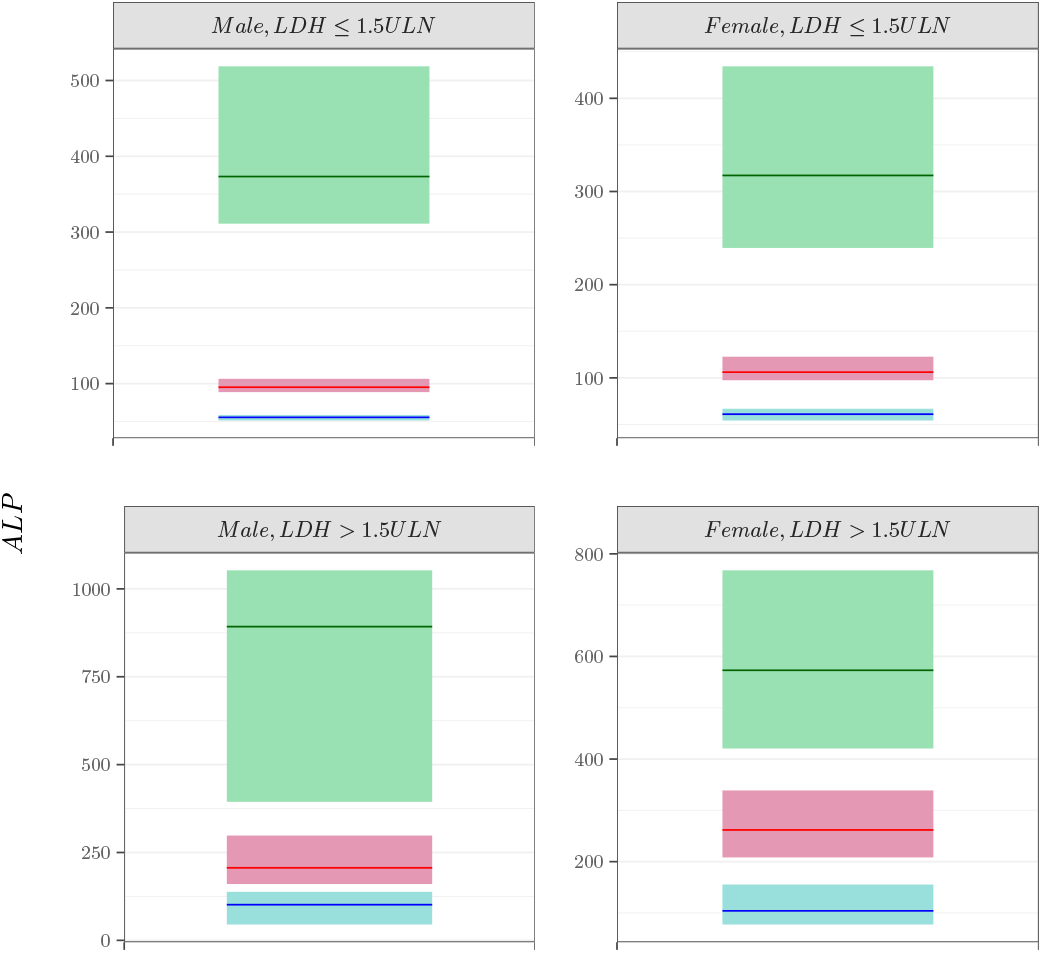
Visual predictive checks of *ALP* density: for each of the 4 covariate combination, the virtual population was simulated 100 times from the copula and the 99% prediction intervals of the percentiles of the distribution were derived and compared to the percentiles observed in the NHANES database. The ribbon areas correspond to the 99% prediction interval of 5^*th*^ (●), 50^*th*^ (●) and 95^*th*^ (●) percentile of the distribution of *ALP*. The lines corresponds to the observed 5^*th*^ (**—**), 50^*th*^ (**—**) and 95^*th*^ (**—**) percentiles.

**TABLE A2.** D-criterion (*ϕ*_*D*_) and its 95% bootsrap confidence interval and runtime for FIM computation using Monte-Carlo and Markov-Chain Monte-Carlo (MCMC) with *n*^(*MC*)^ = 1000 and *n*^(*GLQ*)^ = 5.

| $n^{(MC)}$ | $n^{(GLQ)}$ | $n^{(MCMC)}$ | $\phi_D$ [95% CI] | runtime (h) |
| --- | --- | --- | --- | --- |
| 1000 | 1 | 100 | 72.1 [ 68.6, 74.9] | 0.6 |
| 1000 | 3 | 100 | 104.5 [101.5, 107.0] | 1.7 |
| 1000 | 5 | 100 | 105.8 [103.6, 107.7] | 3.0 |
| 1000 | 7 | 100 | 106.0 [104.1, 107.6] | 4.1 |
| 1000 | 1 | 200 | 71.4 [ 68.4, 73.6] | 0.8 |
| 1000 | 3 | 200 | 104.5 [101.8, 106.4] | 2.2 |
| 1000 | 5 | 200 | 105.8 [103.8, 107.3] | 3.6 |
| 1000 | 7 | 200 | 106.1 [104.5, 107.7] | 5.9 |

**TABLE A3.** Clinical trial simulation with 200 datasets, estimates, relative bias (*RB*), relative root mean squared error (*RRMSE*). * indicates significant bias

| Parameter | Simulation Value | Estimate | $RB$ (%) | $RRMSE$ (%) |
| --- | --- | --- | --- | --- |
| $\mu_w$ | 86.31 | 86.70 | 0.46 | 4.80 |
| $r_{w,LDH}$ | 1.55 | 1.56 | 0.69 | 13.36 |
| $r_{w,ALP}$ (P10) | 0.77 | 0.78 | 0.79* | 3.96 |
| $r_{w,ALP}$ (P90) | 1.69 | 1.67 | -1.14* | 7.93 |
| $\mu_g$ | 0.30 | 0.30 | -0.31 | 5.86 |
| $r_{g,ALP}$ (P10) | 0.84 | 0.85 | 0.75* | 5.10 |
| $r_{g,ALP}$ (P90) | 1.43 | 1.42 | -0.75 | 10.28 |
| $\mu_d$ | 1.79 | 1.80 | 0.55 | 4.72 |
| $\omega_w$ | 0.66 | 0.66 | -0.78* | 4.73 |
| $\omega_g$ | 0.94 | 0.94 | -0.47 | 5.36 |
| $\omega_d$ | 0.67 | 0.67 | -0.13 | 6.44 |
| $b$ | 0.19 | 0.19 | -0.17 | 1.66 |
| $\gamma$ | 3.12 | 3.17 | 1.55* | 7.45 |
| $\beta_0$ | -4.16 | -4.25 | 1.98* | 8.32 |
| $\alpha$ | 4.18 | 4.31 | 3.09* | 10.62 |
| $r_{h,LDH}$ | 2.13 | 2.25 | 5.62* | 26.35 |
| $r_{h,Sex}$ | 0.94 | 0.96 | 1.73 | 16.77 |

**FIGURE A2.**
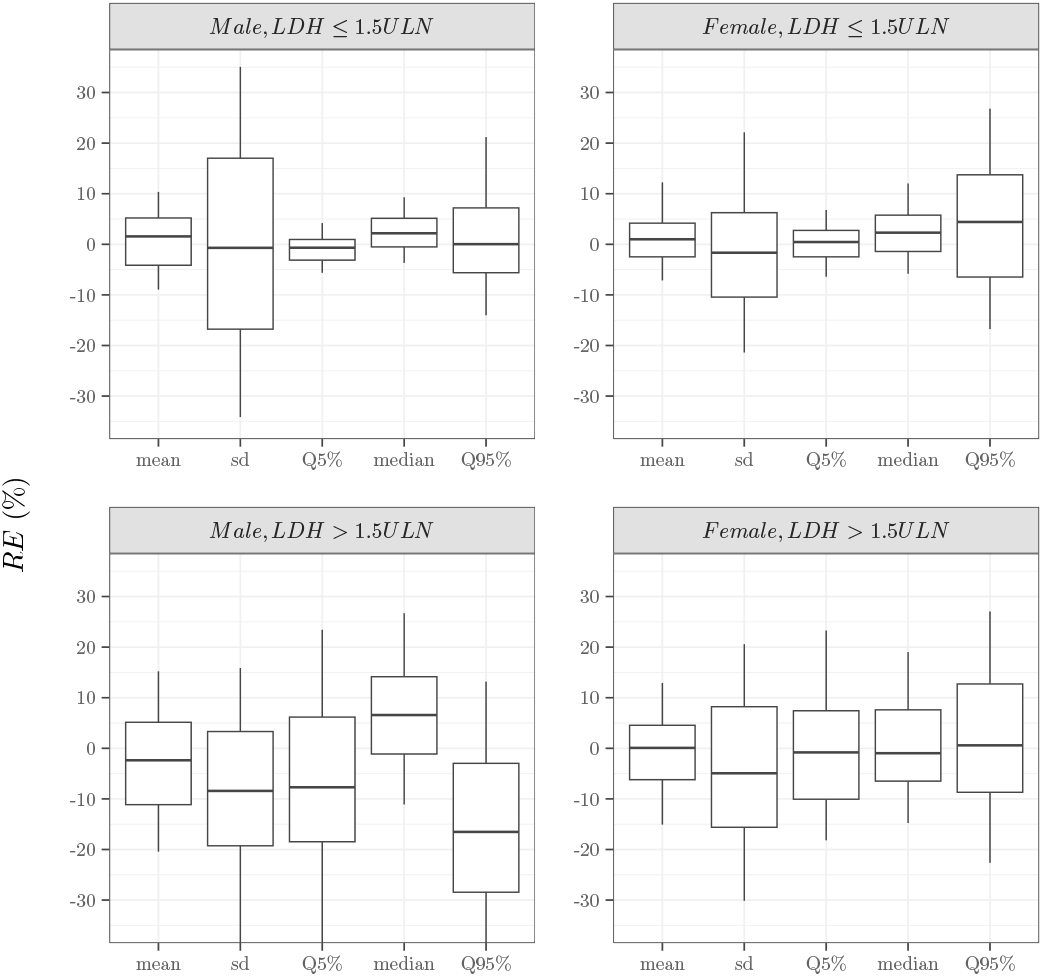
Boxplot of the relative error (*RE*) of in summary statistics (mean, standard deviation and percentiles) of the *ALP* distribution for the 4 covariate combinations, as compared to the statistics of the Horizon database. For each combination, the virtual population was simulated 100 times from the copula. The boxplot displays the median, the 25^*th*^ and 75^*th*^ percentiles, while the whiskers are 5^*th*^ and 95^*th*^ percentiles. sd refers to the standard deviation, and *P*5%, *P*50% and *P*95% to the 5^*th*^, 50^*th*^ and 95^*th*^ percentiles respectively.

**FIGURE A3.**
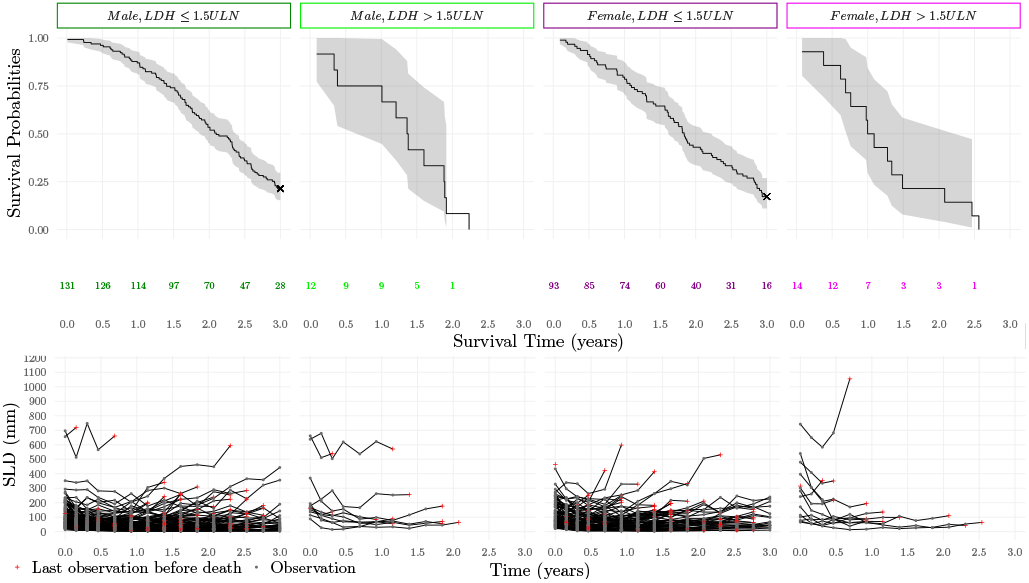
A simulated dataset, Kaplan-Meier curves of survival and longitudinal measurements of sum of lesion diameter (SLD) across the four discrete covariate combinations.

**FIGURE A4.**
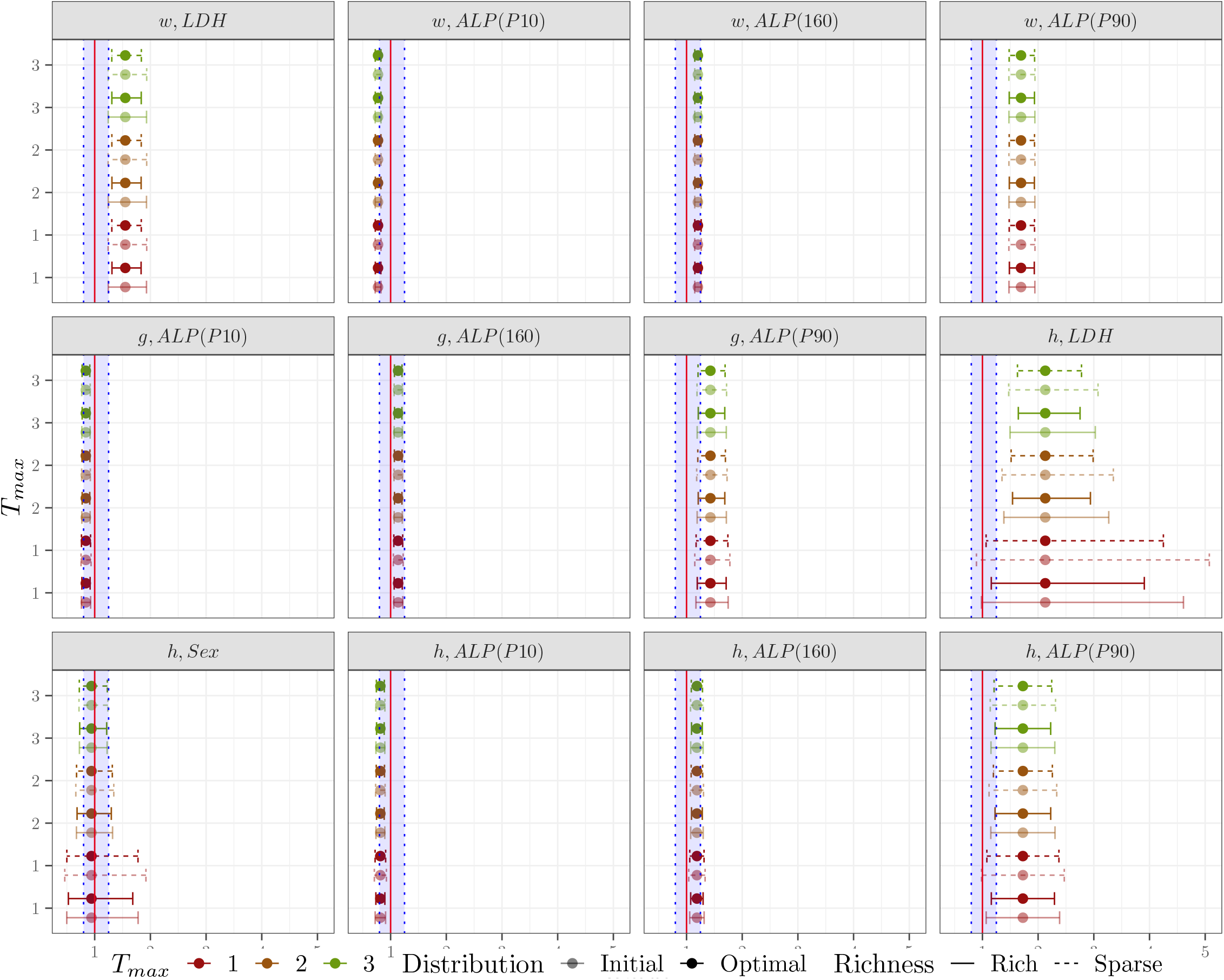
Forest plot obtained with initial distribution vs optimal covariate optimisation for various designs based on Fisher Information Matrix computation using Monte-Carlo and Adaptive Gaussian Quadrature (MC-AGQ) with *n*^(*GLQ*)^ = 5, *n*^(*MC*)^ = 1000 and *n*^(*AGQ*)^ = 5.

## B Appendix 2: censoring of SLD observations

**FIGURE B5.**
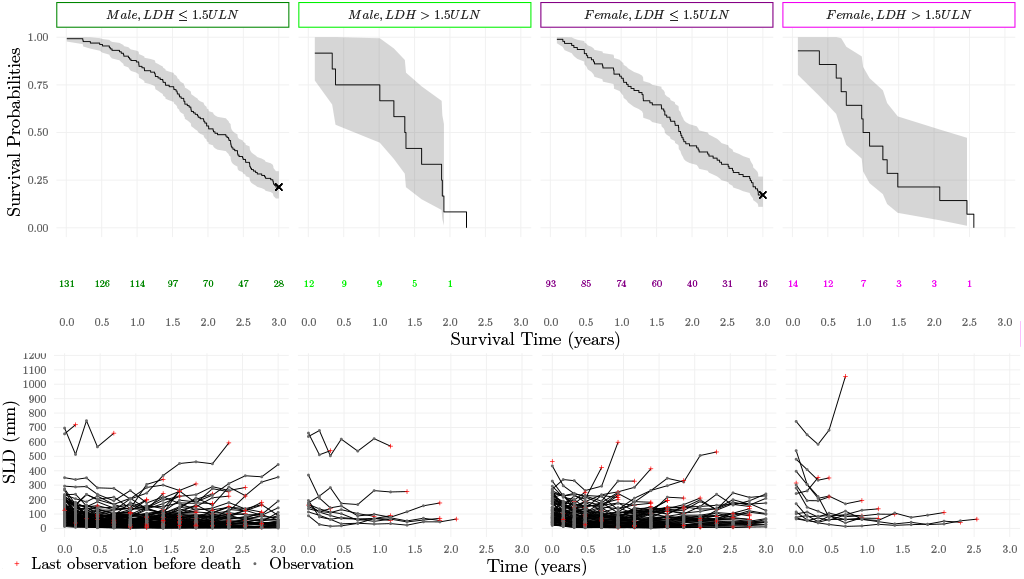
A simulated dataset with sum of lesion diameter (SLD) censoring after disease progression, Kaplan-Meier curves of survival and longitudinal measurements of SLD across the four discrete covariate combinations.

**TABLE B4.** D-criterion (*ϕ*_*D*_) and its 95% bootsrap confidence interval and runtime for Fisher Information Matrix computation using Monte-Carlo and Adaptive Gaussian Quadrature (MC-AGQ) with *n*^(*MC*)^ = 1000 and *n*^(*GLQ*)^ = 7.

| $n^{(MC)}$ | $n^{(GLQ)}$ | $n^{(AGQ)}$ | $\phi_D$ [95% CI] | runtime (h) |
| --- | --- | --- | --- | --- |
| 1000 | 5 | 1 | 88.4 [87.0, 89.6] | 0.2 |
| 1000 | 5 | 3 | 86.6 [85.3, 87.9] | 0.4 |
| 1000 | 5 | 5 | 78.5 [77.2, 79.7] | 1.3 |
| 1000 | 5 | 7 | 76.5 [75.2, 77.6] | 3.4 |

**TABLE B5.** D-criterion (*ϕ*_*D*_) and its 95% bootsrap confidence interval and runtime for FIM computation using Monte-Carlo and Markov-Chain Monte-Carlo (MCMC) with *n*^(*MC*)^ = 1000 and *n*^(*GLQ*)^ = 5.

| $n^{(MC)}$ | $n^{(GLQ)}$ | $n^{(MCMC)}$ | $\phi_D$ [95% CI] | runtime (h) |
| --- | --- | --- | --- | --- |
| 1000 | 5 | 100 | 76.7 [75.2, 78.0] | 5.9 |
| 1000 | 5 | 200 | 76.4 [75.1, 77.6] | 8.2 |

**FIGURE B6.**
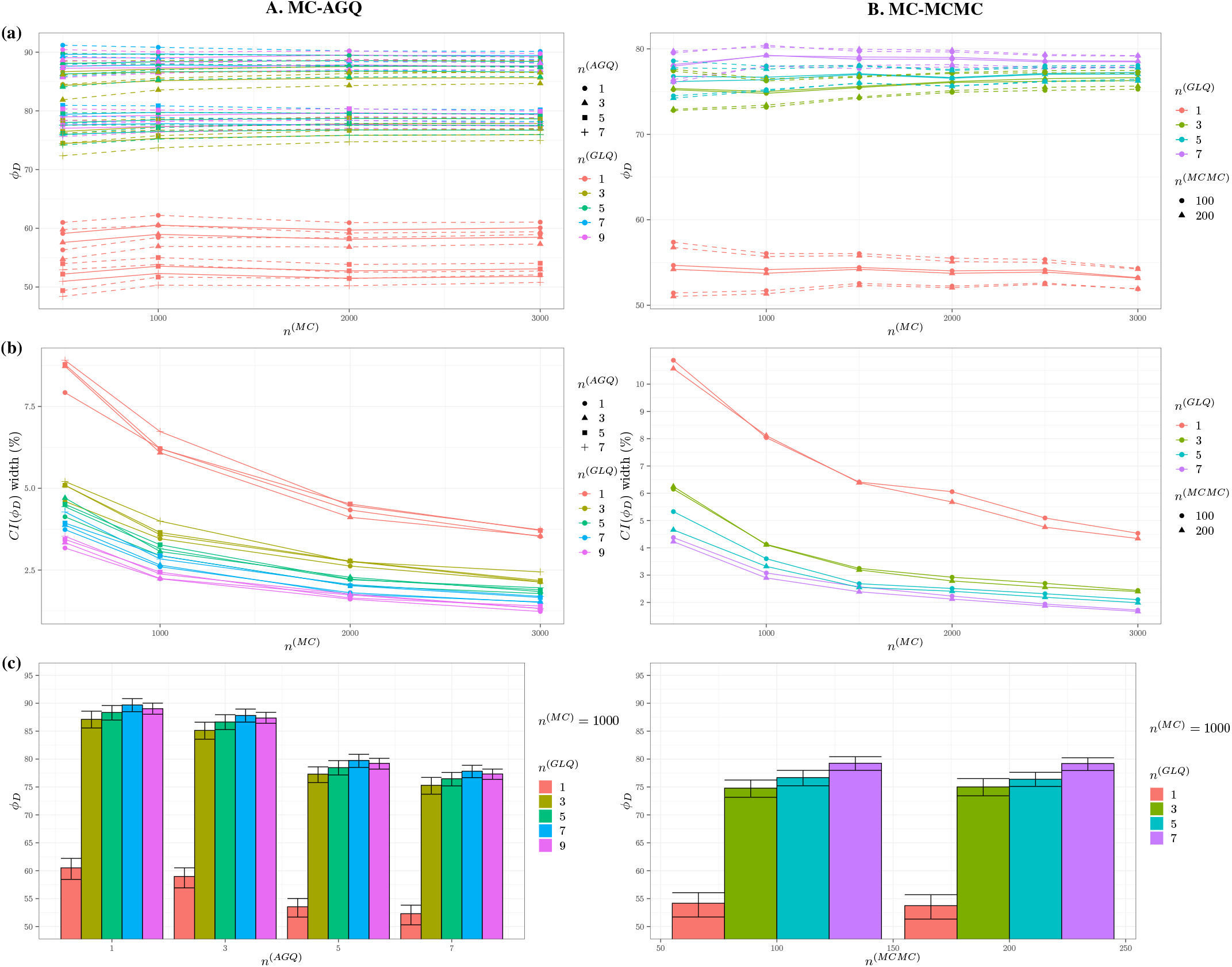
Influence of the hyper-parameters in Fisher Information Matrix computation using Monte-Carlo and Adaptive Gaussian Quadrature (MC-AGQ) on the left, and Monte-Carlo and Markov-Chain Monte-Carlo (MC-MCMC) on the right **(a)** D-criterion (*ϕ*_*D*_) with its 95% confidence interval and **(b)** bootstrap confidence interval on *ϕ*_*D*_ relative width, as a function of the number of Monte-Carlo samples (*n*^(*MC*)^), for increasing number of quadrature nodes in covariate integration (*n*^(*GLQ*)^) and respectively, increasing number of quadrature nodes in random effects integration (*n*^(*AGQ*)^) for AGQ and increasing number of MCMC samples in random effects integration (*n*^(*MCMC*)^) for MCMC **(c)** D-criterion (*ϕ*_*D*_) with its 95% confidence interval with *n*^(*MC*)^ = 1000 and increasing number of quadrature nodes in covariate integration (*n*^(*GLQ*)^) and respectively increasing *n*^(*AGQ*)^ and *n*^(*MCMC*)^.

### B.1 Comparison with CTS

With CTS the empirical D-crtiterion across 200 datasets was 75.14 [74.97; 82.66].

**FIGURE B7.**
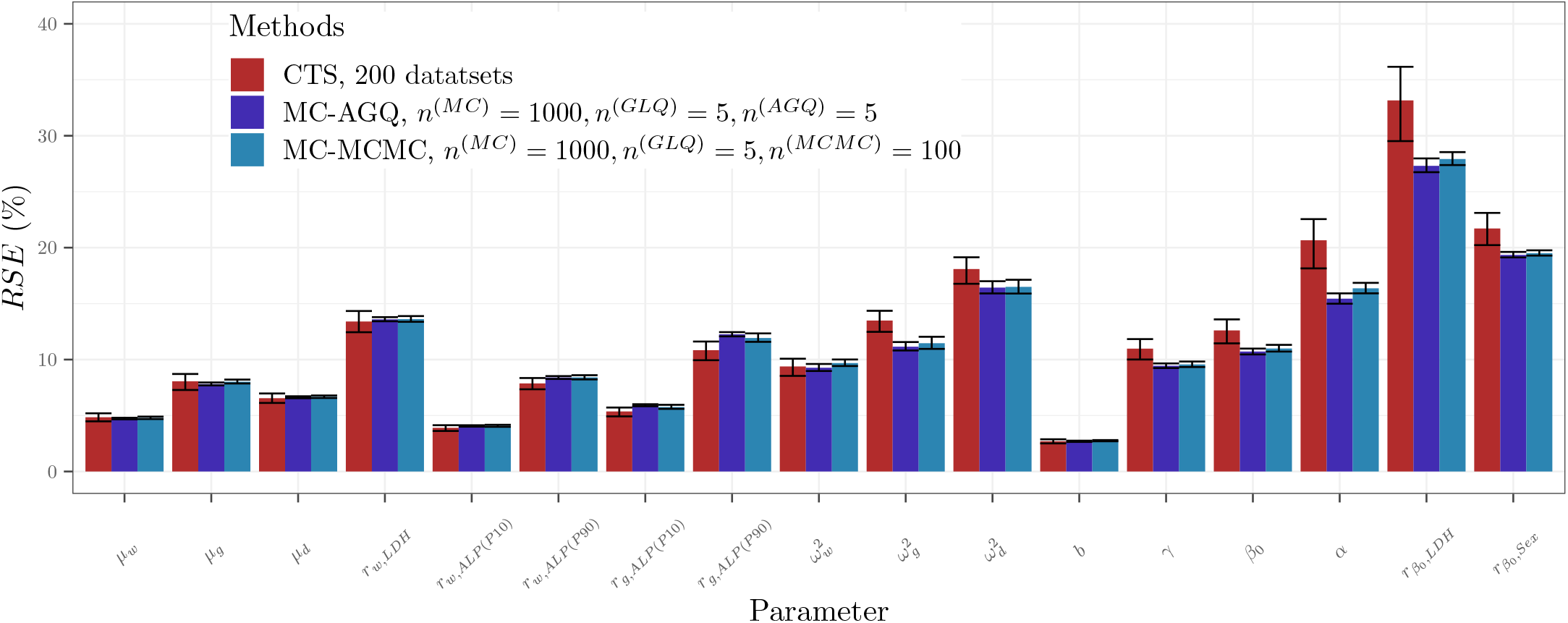
Predicted Relative Standard Errors (RSE) obtained through Clinical trial Simulations (CTS) with 200 datasets, FIM computation using Monte-Carlo and Adaptive Gaussian Quadrature (MC-AGQ) with *n*^(*MC*)^ = 1000, *n*^(*GLQ*)^ = 5, *n*^(*AGQ*)^ = 5, and FIM computation using Monte-Carlo and Markov-Chain Monte-Carlo (MC-MCMC) with *n*^(*MC*)^ = 1000, *n*^(*GLQ*)^ = 5, *n*^(*MCMC*)^ = 100

**TABLE B6.** Clinical trial simulation with 200 datasets, estimates, relative bias (*RB*), relative root mean squared error (*RRMSE*). * indicates significant bias

| Parameter | Simulation Value | Estimate | RB (%) | RRMSE (%) |
| --- | --- | --- | --- | --- |
| $\mu_w$ | 86.31 | 86.72 | 0.48 | 4.86 |
| $r_{w,LDH}$ | 1.55 | 1.56 | 0.73 | 13.39 |
| $r_{w,ALP}(P10)$ | 0.77 | 0.78 | 0.79* | 3.97 |
| $r_{w,ALP}(P90)$ | 1.69 | 1.67 | -1.14* | 7.93 |
| $\mu_g$ | 0.30 | 0.30 | 0.05 | 8.04 |
| $r_{g,ALP}(P10)$ | 0.84 | 0.85 | 0.95* | 5.43 |
| $r_{g,ALP}(P90)$ | 1.43 | 1.41 | -1.05 | 10.86 |
| $\mu_d$ | 1.79 | 1.80 | 0.71 | 6.57 |
| $\omega_w$ | 0.66 | 0.66 | -0.77* | 4.75 |
| $\omega_g$ | 0.94 | 0.93 | -1.09* | 6.84 |
| $\omega_d$ | 0.67 | 0.67 | -0.32 | 9.00 |
| $b$ | 0.19 | 0.19 | -0.01 | 2.70 |
| $\gamma$ | 3.12 | 3.23 | 3.42* | 11.47 |
| $\beta_0$ | -4.16 | -4.35 | 4.37* | 13.31 |
| $\alpha$ | 4.18 | 4.49 | 7.42* | 21.90 |
| $r_{h,LDH}$ | 2.13 | 2.33 | 9.32* | 34.36 |
| $r_{h,Sex}$ | 0.94 | 0.96 | 2.41 | 21.79 |

## Notes

### Competing Interest Statement

The authors have declared no competing interest.

### Author Declarations

The study used only openly available human data that were originally located at: https://data.projectdatasphere.org/projectdatasphere/html/content/78

